# Behaviourally-informed two-way text messaging to improve return to HIV care in South Africa: evidence from a randomised controlled trial

**DOI:** 10.1101/2024.08.19.24312207

**Authors:** Christine Njuguna, Preethi Mistri, Lawrence Long, Candice Chetty-Makkan, Brendan Maughan-Brown, Alison Buttenheim, Laura Schmucker, Sophie Pascoe, Harsha Thirumurthy, Cara O’Connor, Barry Mutasa, Kate Rees

## Abstract

**Introduction:** One-way text messaging to re-engage people in HIV care has shown promise, but little is known about two-way messaging where the recipient is asked to respond. We evaluated a two-way text messaging intervention, informed by behavioural science, to increase re-engagement in care following missed antiretroviral therapy (ART) visits.

**Methods:** We conducted an individual-level randomised controlled trial between February and March 2023 in Capricorn District, South Africa. Adults aged ≥18 years who had missed an ART visit by >28 days were randomised to (1) a standard one-way text message, or (2) behaviourally informed two-way text messages. Two-way messages asked recipients to reply by selecting from a list of reasons for their missed visit. Those who responded received another text message tailored to their response and designed using behavioural economics principles (affect heuristic, availability heuristic, loss aversion, social norms, providing slack, expanding choice). The primary outcome was return to care within 45 days, analysed as 1) intention to treat, and 2) restricted to successful message delivery.

**Results:** 3,695 participants were randomised: 1,845 to the one-way message group and 1,850 to the two-way message group. 27.9% (515/1845) of participants sent a one-way message and 27.2% (503/1850) sent a two-way message returned for an ART visit within 45 days (proportion difference:−0.7%, p-value: 0.622). In an analysis restricted to participants whose text message was delivered, 28.3% (310/1094) in the one-way message group compared to 28.3% (304/1076) in the two-way message group returned to care (proportion difference:−0.09%, p-value: 0.966). 19.5% (210/1076) responded to the two-way message. The two most commonly reported reasons for missed appointments were being out of town (41.0%) and still having medication (31.0%.). Among those who responded, 27.1% (95%CI: 21.3-33.7) returned for an ART visit. Clients ≥50 years were less likely to respond to the two-way text message, (AOR 0.4; 95%CI: 0.2-0.9)

**Conclusions:** Behaviourally informed two-way text messages did not improve return to care over one-way messages. However, they elicited reasons for disengagement, which could inform future outreach for missed visits. Additional research is needed on the mode, content and timing of two-way messages intended to increase return to care.

**Clinical Trial Number:** PACTR202202748760768 & DOH-27-042022-6703.

## Introduction

Retaining people living with HIV (PLHIV) on antiretroviral therapy (ART) remains a challenge in South Africa. In 2023, 95% (7.4 million) of PLHIV knew their HIV status, 75% (5.8 million) of those were accessing ART, and 91% (5.3 million) accessing ART had a viral load of less than 1000 copies/ml [1]. Treatment interruptions are a major barrier to increasing ART coverage and achieving the UNAIDS 95-95-95 targets [2]. Common reasons for disengagement from care include unexpected events/life changes, socio-economic reasons, mobility and lack of perceived benefit of ART [3,4]. In South Africa, loss to follow-up rates among people on ART range from 12-30% [5–8], with concomitant increased HIV transmission, treatment resistance, morbidity, and mortality [9]. In response to increasing disengagement the National Department of Health in South Africa launched the “Welcome Back Campaign” in 2021[10]. Fundamental to this campaign is a client-centred approach that aims to equip personnel with processes to manage treatment interruptions and re-engagements without judgement, and develop key messages for client support [10].

Text messaging in the form of appointment reminders for PLHIV have improved retention in care [11–13]. This intervention can be cost-effective [14] and offers convenience and privacy [15]. Evidence on text messaging for re-engaging PLHIV lost to follow-up is limited. Some studies, primarily from the United States, have reported an improved return to HIV care with the use of one-way text messaging [16,17]. Moreover, there is limited evidence on interventions that have used behavioural economics (BE) principles in the design of text messaging to re-engage PLHIV who are lost to care. Recent results from a study in South Africa using the “fresh start” effect to frame one-way text messages indicated that these messages improved re-engagement in HIV care [18].

Two-way text messaging has shown promise in improving medication adherence [19]. For example, a meta-analysis of randomised controlled trials from low- and middle-income countries found that two-way text messaging was associated with substantially improved medication adherence compared to one-way messaging for hypertension medication, malaria prophylaxis and ART [20]. Two-way text messages may be effective in part because they enable greater interactivity between message recipients and message senders [19,21–23] and an enhanced patient-provider relationship [24].

Health behaviours and decision-making may be influenced by an individual’s experience of poverty and time constraints. Prior research has shown that poverty affects mental bandwidth, attention, and cognitive capacity [25,26]. It can also lead people to focus on immediate needs, thereby reducing the likelihood that they use limited resources for clinic visits [27]. When faced with time constraints due to employment activities or caregiving needs, people experiencing time scarcity may also be highly affected by a lack of flexibility, or slack, in clinic schedules. Separately, PLHIV may also have mental models (internal representations of external reality based on, for example, past experiences and perceptions) that affect their perceptions of whether it is acceptable to return to the clinic after having missed an appointment.

In South Africa, economically disadvantaged groups are less likely to access and utilise healthcare services due in part to the associated financial costs [28]. While access to ART services in the public health system is free in South Africa, ART adherence is impacted through care-related costs like transportation fees to the clinic, purchasing of healthy foods and medications to ease the side effects from ART [29,30]. Competing psychosocial and economic challenges play a key role in ART adherence and treatment interruptions in South Africa [29,31].

Decision-making could also be influenced by two modes of thinking: faster instinctive and emotional thinking which requires less effort, and slower more deliberative consideration requiring reflection and more effort in making decisions or taking action [32,33]. Prompts may be used at a specific moment or in relation to a particular event to motivate decision making [34]. Prompts could also remind a person about something they may have thought about but not acted on to motivate them to decide to act in the moment [34]. Prompts using reason are thought to motivate reflective thinking, and those using emotion, intuitive thinking in making decisions [32]. Based on specific prompts triggering a response, receiving a text message could be considered a cue to further prompt action [35].

This study sought to incorporate BE principles related to conditions of scarcity into two-way messaging, aligned to the Department of Health Welcome Back campaign. The intervention sought to prompt reflective thinking to motivate engagement in care. We compared two-way to one-way text messaging to determine the effects on return to care in PLHIV who had missed ART visits, and add to the limited evidence on two-way messaging for increasing retention in care.

## Methods

### Study design and setting

An individual-level randomised controlled trial (RCT) following the Consolidated Standards of Reporting Trials (CONSORT) guidelines[36] was conducted in Capricorn District, Limpopo Province, South Africa, between February and March 2023. Capricorn District has a population of 1.3 million and HIV prevalence among people aged 15-49 years is 8.1% [37]. A tracing programme outlined in the national ART guidelines aims to support PLHIV to return to care after disengagement [38]. Tracing usually consists of three telephonic attempts and one home visit attempt if telephonic tracing is unsuccessful. If the home visit is unsuccessful, the client is considered lost from care [38].

### Study population

We extracted data from TIER.Net, a national electronic database comprising demographic and clinical data for people on ART [39]. Eligibility criteria included those 18 years or older, having missed an ART appointment by more than 28 days, and with a 10-digit cell phone number on record. We excluded clients who had been documented as deceased or transferred to another clinic.

### Randomisation

Eligible clients were randomised using a 1:1 ratio into: 1) a one-way text message group; and 2) a BE-informed two-way text message group. Computer-generated numbers were used to generate a random allocation sequence.

### One-way text message group

In addition to routine tracing, this group received the following standard of care one-way text message encouraging return to the clinic, aligned with a national campaign for a return to care (i.e., the “Welcome Back Campaign”)[3,40] : “*Hi, this is your clinic. We noticed you missed an appointment. We invite you to return soon. Check in at Reception, we are ready to help. You are welcome back!”* The readability (Flesch Kincaid test) [41] of the text message was 5^th^-grade level (very easy to read). All text messages were developed by a panel of ART programme and BE experts and delivered by Anova Health Institute, a PEPFAR/USAID implementing partner, using a bulk text messaging service.

### BE-informed two-way text message group

In addition to routine tracing, this group received a BE-informed two-way text message that asked about reasons for missing a clinic appointment. The text message read: *Hi, this is your clinic. Could you share why you missed your appointment? Reply 1: Out of town 2: Too busy 3: Still have meds 4: Clinic not friendly 5: Other reason. Free to reply.* The readability of the text message was 6^th^-grade level (easy to read).

For those who replied, a second message was sent, tailored to the reason for their missed appointment. For example, if the system received a reply of “1” or “out of town”, the second message read: “*You can go to any nearby clinic to get your medication, even without a transfer letter. No one should ever be turned away from a clinic. You can also ask for extra medication if you are going away. You are welcome back!”* We limited the length to 160 characters to ensure delivery as a single text message. Also, since most South African mobile users are charged to send text messages, we provided reverse billing, removing this cost to the sender and noted this in the message.

The two-way text message content was developed using BE principles to encourage return to care (**Table 1**). Specifically, we leveraged cognitive biases related to past experiences to prompt reflective thinking and shape decision-making. Messages were designed to provide greater slack by introducing more flexibility in returning to care, for example by telling participants they could collect medication at any clinic even without a transfer letter or they could collect medication at more convenient locations. Messages were also framed to address mental models around the clinic being unfriendly. Finally, messages leveraged loss aversion (a real or potential loss is perceived by individuals as psychologically or emotionally more severe than an equivalent gain [42]), the affect heuristic (representations of events in people’s minds are tagged with emotion from past experiences [43,44]), and social norms (informal rules of beliefs, attitudes, and behaviours that are considered acceptable in a particular society or social group [45]).

**Table 1:** Standard of care and intervention text messages including tailored responses.

| SOC Message | Intervention Message | Response text messages to recipient responses to intervention text messages |
| --- | --- | --- |
| “Hi, this is your clinic. We noticed you missed an appointment. We invite you to return soon. Check in at Reception, we are ready to help. You are welcome back!” | “Hi, this is your clinic. Could you share why you missed your appointment? Reply<br>1: Out of town<br>2: Too busy<br>3: Still have meds<br>4: Clinic not friendly<br>5: Other reason<br>Free to reply.” | 1. Out of town:<br>“You can go to any nearby clinic to get your medication, even without a transfer letter. No one should ever be turned away from a clinic. You can also ask for extra medication if you are going away. You are welcome back!”<br>2. Too Busy:<br>“Ask your clinic about easy options for collecting your meds. It might be possible to pick up meds at Clicks, the Post Office or a pharmacy closer to you. You are welcome back!”<br>3. Still have meds:<br>“Please come back to the clinic before you run out of meds. Don't lose your chance to stay healthy. Take your meds every day. You are welcome back!”<br>4. Clinic not friendly:<br>“It can be scary to come back to the clinic. But it is important for your health. Our clinic is working to be friendly. You are welcome back!”<br>5. Other reason:<br>“Don’t worry that you have missed your appointment. These things happen to everyone! Make time to visit the clinic soon. You are welcome back!”<br>No response after 3 days<br>“Don’t worry that you have missed your appointment. These things happen to everyone! Make time to visit the clinic soon. You are welcome back!” |

### Sample size calculation

We included clients who had missed an appointment by >28 days and <180 days. The six-month cut-off was to prevent contamination from prior studies. Based on preliminary data from a study on text messaging to improve return to care [18], a return rate for the one-way text message group in our study was assumed to be 30%. Using a significance level of 0.05, power of 0.8, and 4% difference in the primary outcome between study groups, we estimated a minimum sample size in each group of 1677 clients (3354 total). As the available sample frame was 4500, we included all eligible participants to increase the precision of estimates.

### Primary outcome

The primary outcome was a binary indicator of whether the participant returned for an ART clinic visit within 45 days of the first text message.

### Statistical analysis

We compared participant characteristics between the study groups and assessed differences using Chi-squared tests. The primary analysis was an intention-to-treat analysis, which included all participants sent a text message, measuring the difference in return to care between the two study groups. The secondary analysis was a per-protocol analysis, restricted to the subset of participants whose text message was delivered (according to the bulk SMS platform), measuring the difference in return to care between the two study groups. To explore the characteristics of those who responded to the two-way message (a key factor in the success of the intervention) we used multivariable logistic regression to assess the association between response and the following variables: age at randomisation, gender, enrolment in differentiated models of care, treatment interruption stratification, priority clinic (defined as high volume ART clinics), ART duration, and sub-district/location. Variables were selected *a priori* based on their availability in TIER.Net. Analyses were conducted using STATA 18 (StataCorp, College Station, Texas).

### Ethical approval

Ethical approval was provided by the University of Witwatersrand Human Research Ethics Committee (HREC) (220207), the University of Pennsylvania Institutional Review Board (IRB) (851123), Boston University IRB (H-42789), and the Limpopo Province Research Committee. No written informed consent was obtained, as per a waiver granted by the HREC.

## Results

In total, 4500 participants met the inclusion criteria and were randomised to the two study groups (2250 in each arm). When ascertaining outcomes, 18.0% (405/2250) and 17.8% (400/2250) were excluded from the one-way message and two-way message groups, respectively, for the following reasons: registered as deceased (22/4500, 0.5%), transferred to another clinic (90/4500, 2%), did not miss an appointment by more than 28 days (251/4500, 5.6%), or they returned before randomization/before text message was sent (442/4500, 9.8%) These misclassifications were due to data capturing backlogs. The final analysis included 1845 participants from the one-way message group and 1850 participants from the two-way message group **(Figure 1**). Most participants were female (65.0%), between the ages of 25-49 years (73.4%), and had been on ART for >12 months (76.5%). Participant characteristics did not differ by study group. **(Table 2)**

**Figure 1:**
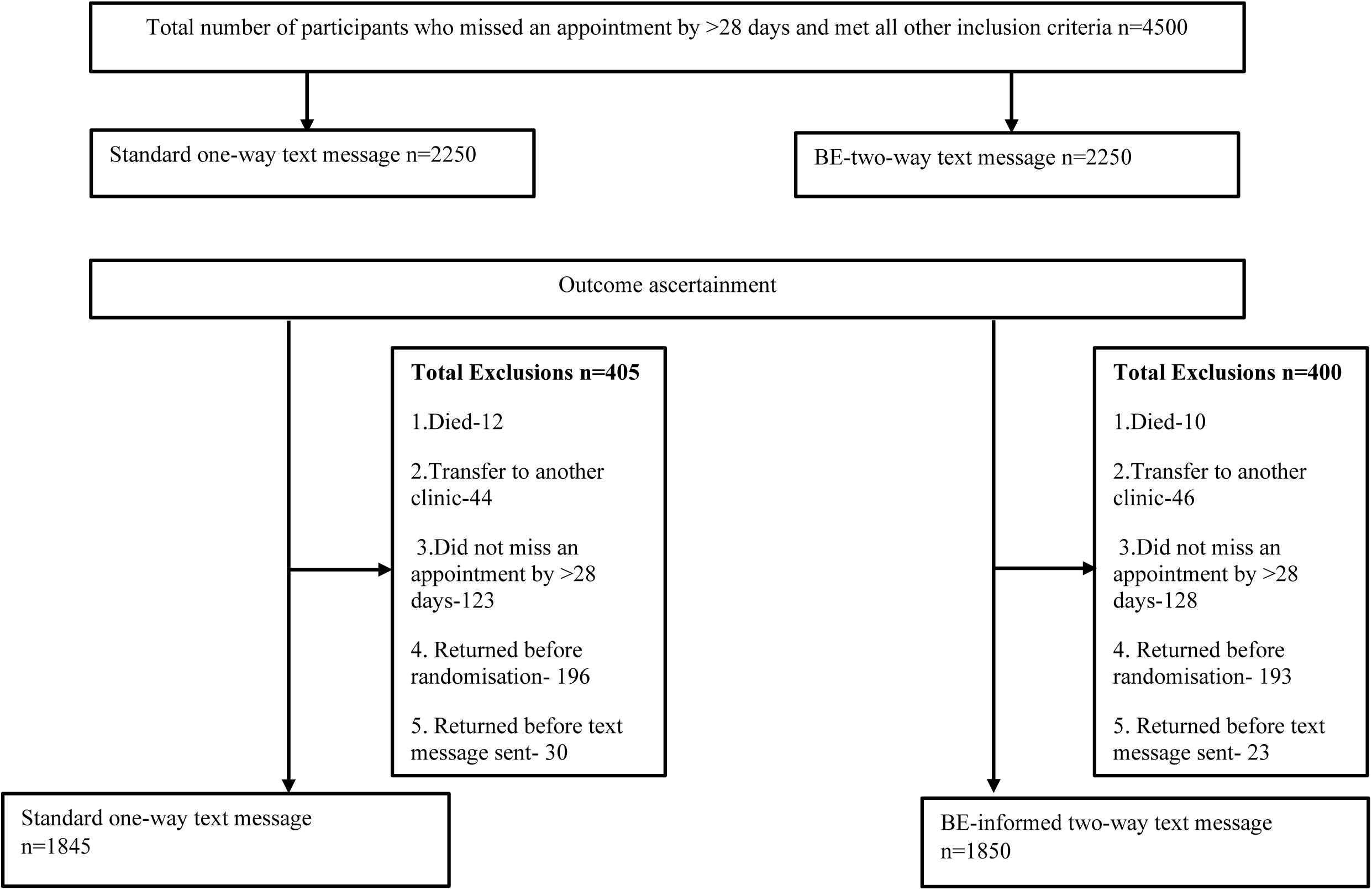
Study consort.

**Table 2:** Characteristics of participants randomised to one-way text message group and BE-informed two-way text message group.

| **Participant characteristics** | **One-way text message** |  | **BE-informed two-way text message** |  |
| --- | --- | --- | --- | --- |
| **Age at randomization (years)** | **N=1845** | **Percent (%)** | **N=1850** | **Percent (%)** |
| 18-24 | 138 | 7.48 | 151 | 8.16 |
| 25-49 | 1348 | 73.06 | 1365 | 73.78 |
| ≥50 | 359 | 19.46 | 334 | 18.05 |
| **Gender** |  |  |  |  |
| Female | 1219 | 66.07 | 1183 | 63.95 |
| Male | 626 | 33.93 | 667 | 36.05 |
| **ART duration (months)** |  |  |  |  |
| <6 | 263 | 14.25 | 275 | 14.86 |
| 6 to12 | 176 | 9.54 | 153 | 8.27 |
| >12 | 1406 | 76.21 | 1422 | 76.86 |
| **Treatment interruption stratification (months)** |  |  |  |  |
| <3 | 1030 | 55.83 | 1096 | 59.24 |
| ≥3 | 815 | 44.17 | 754 | 40.76 |
| **Differentiated models of care** |  |  |  |  |
| Yes | 660 | 35.77 | 696 | 37.62 |
| No | 1185 | 64.23 | 1154 | 62.38 |
| **Priority clinic** |  |  |  |  |
| No | 820 | 44.44 | 823 | 44.49 |
| Yes | 1025 | 55.56 | 1027 | 55.51 |
| **Sub-district** |  |  |  |  |
| Blouberg | 256 | 13.88 | 217 | 11.73 |
| Lepelle-Nkumpi | 259 | 14.04 | 264 | 14.27 |
| Molemole | 170 | 9.21 | 201 | 10.86 |
| Polokwane | 1160 | 62.87 | 1168 | 63.14 |

### Return to care

#### Text message delivery rates

99.9% (3691/3695) of text messages were sent. Only four had invalid cell phone numbers. Overall, 58.8% (2170/3691) of text messages were recorded as having been delivered. Delivery rates were similar in the two groups, 59.4% (1094/1842) in the one-way message group and 58.2% (1076/1849) in the two-way message group.

#### Intention to treat analysis

There was no difference in the primary outcome between groups with just over a quarter of participants re-engaging in care after the messages were sent: 27.9% (515/1845; 95% CI: 28.9-30.0) in the one-way message group and 27.2% (503/1850; 95% CI: 25.2-29.3) in the two-way message group (proportion difference=−0.7%; p-value 0.622). **(Table 3)**

**Table 3:** Return to care at 45 days for participants randomised to one-way text message group and the two-way BE-informed text message group.

| Text message group | Sample size | Number of participants returning to care | Return to care & 95% CI | % difference | P-value |
| --- | --- | --- | --- | --- | --- |
| **All participants- Intention to treat analysis** |  |  |  |  |  |
| Standard one-way text message | 1845 | 515 | 27.91% (28.88-30.02) | 0.72% | 0.622 |
| BE-informed two-way text message | 1850 | 503 | 27.19% (25.17-29.28) |  |  |
| **Participants with text messages delivered- Per protocol analysis** |  |  |  |  |  |
| Standard one-way text message | 1094 | 310 | 28.34% (25.68-31.11) | 0.09% | 0.966 |
| BE-informed two-way text message | 1076 | 304 | 28.25% (25.58-31.05) |  |  |
| **Participants who responded to the two-way text message** |  |  |  |  |  |
| Yes | 210 | 57 | 27.14% (21.25-33.69) | 1.38% | 0.690 |
| No | 866 | 247 | 28.52% (25.53-31.66) |  |  |

#### Per-protocol analysis

When restricting the analysis to those participants whose message was delivered, the main findings remained unchanged with just over a quarter of participants re-engaging in care; 28.3% (310/1094; 95%CI: 25.7-31.1) in the one-way message group and 28.3% (304/1076; 95%CI: 25.6-31.1) in the two-way message group (proportion difference=0.1%, p-value 0.966). **(Table 3)** In all analyses, re-engagement in ART care did not differ by study group when stratified by duration of treatment interruption (<3 months and ≥3 months). (results not reported).

### Reasons for missing appointments

In the two-way message group, 19.5% (210/1076) responded to the message. There were 210 clients who responded to the two-way message with a total of 238 responses. Twenty-eight clients responded with more than one reason. The reported reasons for missing appointments were: out of town 41.0% (86/210), too busy 15.2% (32/210), had medication 31.0% (65/210), clinic unfriendly 7.6% (16/210), other 18.6% (39/210). **(Figure 2)**

**Figure 2:**
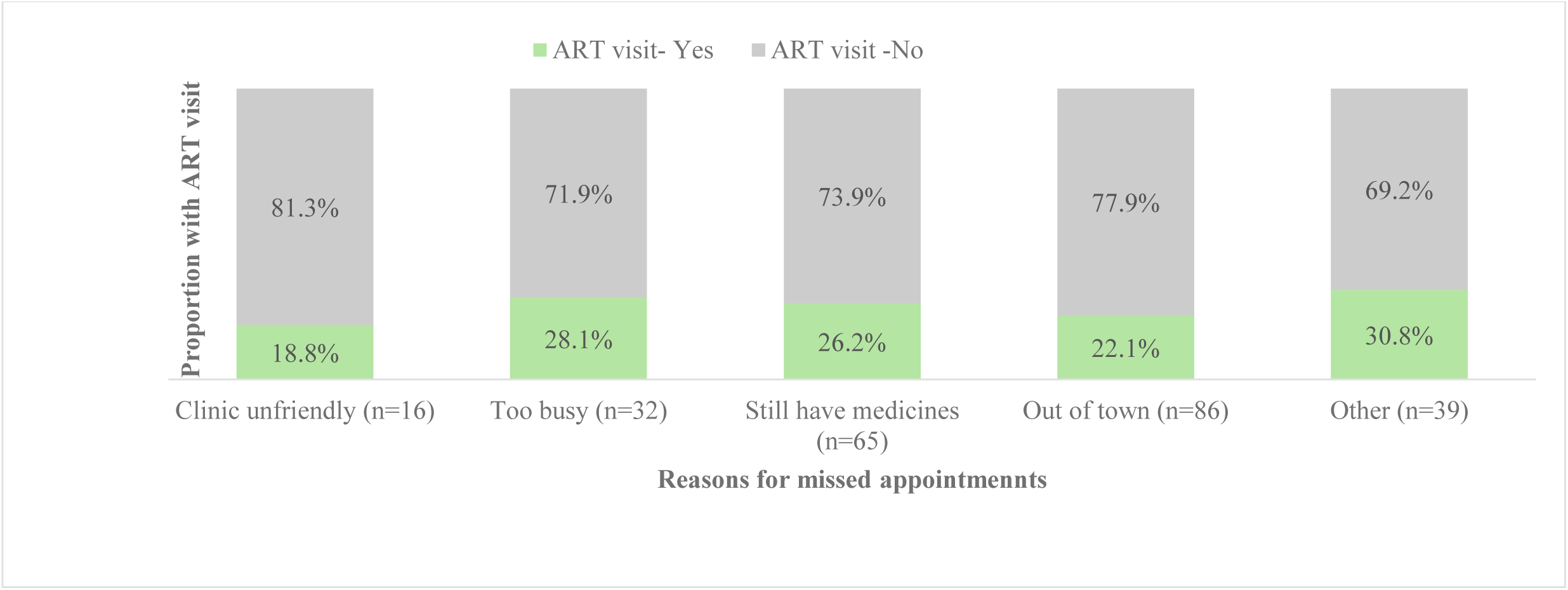
Reasons for missed appointments and ART visit proportions by reasons for missed appointment (N=210). †A client should select more than one category.

The difference in return to care within 45 days between participants who responded to the text message in the two-way message group (n=210) (27.1%, 95% CI: 21.3-33.7) and participants in the two-way message group who did not respond to the message (28.5%; 95%: 25.5-31.7) was not statistically significant. (**Table 3)** Stratifying by reason for missing an appointment, participants who responded, “clinic not friendly” returned for an ART visit less often (18.8%) compared to 30.8% of those who responded “other’’ and 28.1% who responded, “too busy” (**Figure 2**).

Multivariable logistic regression to explore factors associated with response to the two-way text message **(Table 4)** found participants ≥ 50 years old (AOR 0.4; 95%CI:0.2-0.9) were less likely to respond compared to those 18-24 years old. Participants enrolled in clinics from Molemole (AOR 2.4; 95%CI:1.2-4.9) and Polokwane (AOR 2.2; 95%CI:1.2-4.1) were more likely to respond compared to those enrolled from Blouberg sub-district.

**Table 4:** Multivariable logistic regression of association between response to two-way text message and participant characteristics (N=1076)

| Participant variables | Responded -<br>Yes (n=210) | Did not respond<br>(n=866) | Unadjusted odds<br>ratio | 95% CI | P-value | Adjusted odds<br>ratio | 95% CI | P-value |
| --- | --- | --- | --- | --- | --- | --- | --- | --- |
| **Age at randomisation**<br>**(years)** |  |  |  |  |  |  |  |  |
| 18-24 | 16 (22.22%) | 56 (77.78%) | 1 |  |  | 1 |  |  |
| 25-49 | 172 (21.37%) | 633 (78.63%) | 0.95 | 0.53-1.70 | 0.865 | 0.93 | 0.51-1.69 | 0.807 |
| >=50 | 22 (11.06%) | 177 (88.94%) | 0.44 | 0.21-0.89 | 0.022 | 0.43 | 0.20-0.89 | 0.024 |
| **Gender** |  |  |  |  |  |  |  |  |
| Female | 148 (21.26%) | 548 (78.74%) | 1 |  |  | 1 |  |  |
| Male | 62 (16.32%) | 318 (83.68%) | 0.72 | 0.52-1.00 | 0.051 | 0.77 | 0.55-1.08 | 0.137 |
| **ART duration (months)** |  |  |  |  |  |  |  |  |
| <6 | 29 (15.93%) | 153 (84.07%) | 1 |  |  | 1 |  |  |
| 6 to12 | 16 (18.82%) | 69 (81.18%) | 1.22 | 0.62-2.40 | 0.557 | 1.32 | 0.66-2.62 | 0.428 |
| >12 | 165 (20.40%) | 644 (79.60%) | 1.35 | 0.88-2.08 | 0.172 | 1.40 | 0.90-2.19 | 0.139 |
| **Treatment interruption**<br>**(months)** |  |  |  |  |  |  |  |  |
| <3 | 138 (20.91%) | 522 (79.09%) | 1 |  |  | 1 |  |  |
| ≥3 | 72 (17.31%) | 344 (82.69%) | 0.79 | 0.58-1.09 | 0.147 | 0.78 | 0.56-1.07 | 0.121 |
| **Priority clinic** |  |  |  |  |  |  |  |  |
| No | 80 (16.16%) | 415 (83.84%) | 1 |  |  | 1 |  |  |
| Yes | 130 (22.38%) | 451 (77.62%) | 1.50 | 1.10-2.04 | 0.011 | 1.31 | 0.94-1.84 | 0.113 |
| **Sub-district** |  |  |  |  |  |  |  |  |
| Blouberg | 13 (10.66%) | 109 (89.34%) | 1 |  |  | 1 |  |  |
| Lepelle-Nkumpi | 21 (14.09%) | 128 (85.91%) | 1.38 | 0.66-2.88 | 0.397 | 1.45 | 0.69-3.05 | 0.327 |
| Molemole | 28 (22.58%) | 96 (77.42%) | 2.45 | 1.20-4.99 | 0.014 | 2.39 | 1.16-4.90 | 0.018 |
| Polokwane | 148 (21.73%) | 533 (78.27%) | 2.33 | 1.27-4.26 | 0.006 | 2.20 | 1.18-4.11 | 0.013 |

## Discussion

Novel, low-cost and scalable interventions are needed to increase re-engagement in HIV care in lower and middle-income countries. Our evaluation of a two-way text messaging intervention incorporating BE principles found no difference in return to care between one-way and two-way messages (even when restricted to successful message delivery). However, the two-way messages were able to elicit reasons for disengagement which could assist programmes in their tracing efforts.

While results have been mixed, previous studies show that text messaging in general can be effective. One-way text messages framed using BE principles (such as *salience, fresh start effect*) have been used to improve ART adherence [46], linkage to ART [47], and re-engagement [18]. Two-way text messaging (without explicit application of BE) has shown promise in HIV care in improving medication adherence [20] and retention, when compared to no text messaging [48]. BE principles in two-way text messaging (*clinician endorsement, endowment effect)* have previously been used to successfully improve COVID-19 vaccine uptake [49]. However, fewer studies have directly compared one-way and two-way text messaging. One study in Kenya using two-way text messages for postpartum women reported similar results to our study, with no significant differences in clinic visit attendance, viral suppression, or adherence between two-way and one-way text messaging [50]. There is potential that both messaging interventions in our study had an impact (above what may have happened in a no message group) but the difference between the groups (one-way and two-way text message) was small.

The similar return rates in our two study groups may be explained in part by the low response to the two-way text message (19.5%) and only sending the two-way text message once. Other studies measuring the effectiveness of continuous two-way text messaging in HIV care and prevention have reported response rates ranging from 49-57% [51–53], although improved outcomes (clinic visits) were noted amongst pre-exposure prophylaxis (PrEP) users only [53]. Our response rates were lower. For care recipients who did not respond to the two-way message, the opportunity for exposure to the BE-tailored message was lost. A possible reason for the low response to the first text message is that people did not understand that replying to the text message was free. It is also possible that the two-way text message was perceived as judgemental since it assumed and asked why the client had missed an appointment. Qualitative studies would provide valuable insights on reasons for non-response to the two-way text message. To increase response rates in future studies, other more flexible platforms such as WhatsApp could be considered to allow easy tracking of conversation threads and longer messages.

Despite the lack of difference between study groups in return to care, our study was able to elicit reasons for missed appointments with the response “out of town” and “too busy” combined accounting for 56% of responses. This finding aligns with a recent systematic review examining reasons for disengagement from HIV care which described acute or unexpected events as significant precipitators of treatment interruptions [4]. These events included unplanned travel, and unexpected social commitments (including caring for others, family or social responsibilities) [4]. PLHIV disengaging from care due to migration or travel and time constraints has been documented in previous studies [3,54–56] with reasons for treatment interruptions often overlapping and being triggered by unexpected events and life changes [4]. These findings underscore the importance of patient-centred care, and the need to ensure accessibility of interventions such as multi-month dispensing or other differentiated service delivery models to improve retention. A third of clients reported still having medicines as a reason for missing appointments. Another study from Malawi reported having medication as a reason for missed appointments, with clients reporting that they had obtained ART from other sources such as friends or other clinics [56]. In our setting, clinic appointments may be scheduled with the intention that the client may have seven days of medication in hand at the time of the appointment. Over time, this supply accumulates. Inconsistencies in medical records and data capturing could also contribute. This highlights the importance of optimising tracing procedures and effective rescheduling to minimise wasted resources and the burden of unnecessary communication for clients. Two-way text messages or chatbots could play a role in tracing procedures if response rates can be improved. Another potential solution is sending further text messages that would enable the client to reschedule their clinic appointment based on the amount of medication.

There were substantial differences in the proportions returning to care across different reasons for missing an appointment. Previous research [3] has shown that people miss appointments because of life circumstances and return to care when it is convenient. While a minority of participants (7.6%) responded “clinic unfriendly” as a reason for missing a clinic appointment, this is likely to be underreported (as the message was sent from the clinic) and return to care was lowest among people who responded that the clinic was unfriendly. This indicates the importance of promoting a friendly, non-judgemental approach that encourages the re-integration of those returning to care, as outlined in national guidelines. It also suggests that further research is warranted to assess the relative role of perceived clinic friendliness as a factor influencing the effectiveness of interventions to encourage re-engagement.

Results from our study indicate that response rates to two-way messaging interventions may vary across populations, suggesting the need for tailoring of messaging to client demographic characteristics. Findings indicate that older individuals may be less likely to respond to the two-way text message, which aligns with a study on texting dependency showing decreased texting with increasing age [57]. Our results also showed higher responses rates in certain sub-districts (Molemole and Polokwane), which may indicate differences by geographic context.

The strengths of this study were the randomised controlled trial design and use of routine electronic data. Limitations included delays in capturing data on TIER.Net, which meant some data determining eligibility were incomplete at the time of extraction. This resulted in some ineligible participants (primarily those who had not missed an appointment by >28 days) who later had to be excluded, reducing the sample size. Secondly, we relied solely on electronic data for ascertainment of return to care without medical record verification; therefore, our estimates may be an underestimate. These findings are likely to be generalisable to similar settings, however, context-specific elements like the cost of mobile services and language are likely to impact on effectiveness.

## Conclusions

Behaviourally informed two-way text messages did not improve return to care. However, among those who responded, the intervention elicited information on reasons for missed visits, which could inform future outreach and provide an opportunity to alter the perception of clinics as non-welcoming. Future research is needed on the mode, content and timing of two-way messages intended to increase return to care.

## Authors’ contributions

CN, LL, PM, CC-M, BM-B, AB, LS, KR conceptualised the study. BM contributed to data curation. CC-M, PM and KR supervised study implementation and data analysis. CN conducted the data analysis. All authors contributed to manuscript writing and approved the final manuscript.

## Data Availability

All data produced in the present study are available upon reasonable request to the authors.

## Acknowledgements

We would like to acknowledge the Limpopo Department of Health, including health clinic staff, and the Anova staff who managed the study procedures, in particular Basani Maluleke and Ntsetse Kgopong.

## Data availability statement

Data sharing for the participant characteristics and the primary outcome of the study (return to care) is not applicable as this data was extracted from the National Department of Health public health programme system (TIER.net) for this analysis (Table 1). Data on text message delivery rates and message responses (Table 2) are available from the corresponding author upon reasonable request.

## List of abbreviations

ART: antiretroviral therapy.
BE: behavioural economics.
HIV: human immunodeficiency virus.
PLHIV: people living with HIV.
RCT: randomised controlled trial.

